# Robust human genetic evidence supporting causal effects of FGF21 on reducing alcohol consuming behaviours

**DOI:** 10.1101/2025.11.15.25340305

**Authors:** Héléne T. Cronjé, Sile Hu, Rachel Gurrell, Marijana Vujkovic, Susanna C. Larsson, Richard Mason, Richard Butt, Dipender Gill

## Abstract

**Introduction:** Alcohol use disorder (AUD) represents a tremendous societal burden, yet few efficacious therapies are available and widely used. Pre-clinical and human observational data support fibroblast growth factor 21 (FGF21) as a promising therapeutic target for the treatment of AUD. Here, we identify a robust genetic instrument for FGF21 agonism and leverage it in the Mendelian randomization paradigm to explore the effects of FGF21 agonism on AUD and related traits, as well as metabolic outcomes more widely.

**Methods:** We first compared associations with the positive control outcomes of liver fat and liver cirrhosis risk for the FGF21 cis-protein quantitative trait Iocus (cis-pQTL) (rs838131) to those for the common allele FGF21 L174P missense variant (rs739320). Having identified the L174P missense variant as a clinically validated genetic instrument, we subsequently performed unweighted Mendelian randomization investigating effects on AUD, related traits, and metabolic outcomes more widely. Finally, we performed colocalization analyses to test whether observed association results reflect a causal mechanism that overlaps with the clinical effects of FGF21 on liver fat and liver cirrhosis.

**Results:** Consistent Mendelian randomization and colocalization evidence support a protective association between genetically predicted FGF21 agonism and alcohol consumption (Mendelian randomization p=1x10^-18^, colocalization posterior probability=0.90), problematic alcohol use (Mendelian randomization p=0.02, posterior probability=0.64), and AUD (Mendelian randomization p=9x10^-8^, posterior probability=0.97). Similar evidence was also observed for favourable effects of FGF21 on improving kidney function, lowering triglyceride levels, lowering proportional energy intake from carbohydrates, increasing proportional energy intake from protein and fat, increasing body weight and lowering waist-to-hip ratio.

**Conclusion:** This study identifies a clinically validated genetic instrument for FGF21 effects to provide robust causal human evidence supporting favourable effects of FGF21 analogues for the treatment of AUD and related traits, as well as on metabolic outcomes more broadly. Further clinical study is duly warranted.

## Introduction

Fibroblast growth factor 21 (FGF21) is a circulating hormone that is primarily produced in the liver and has been implicated in glucose and lipid metabolism.^1^ Despite initially being pursued as a therapeutic target in diabetes and obesity with limited success,^2^ FGF21 analogues have demonstrated promising signs of efficacy for treating severe hypertriglyceridemia,^3^ metabolic dysfunction-associated steatohepatitis (MASH) and liver cirrhosis.^4^

Given the pleiotropic effects of FGF21,^5^ its analogues may also offer additional therapeutic opportunities. Specifically, pre-clinical studies have supported effects of FGF21 on broadly suppressing alcohol consumption behaviours through mechanisms that affect alcohol palatability and modulation of neuronal activity in the nucleus accumbens.^6^ Such data have highlighted FGF21 analogues as a potential therapy for treating alcohol use disorder (AUD).^7^ With 7% of the global population aged over 15 years estimated to have lived with AUD,^8^ and few efficacious and safe treatments currently available,^9^ developing novel therapies remains a research and development priority.

Here, we employ the Mendelian randomization paradigm by leveraging large-scale human genetic data to explore the causal evidence for effects of FGF21 on alcohol consumption behaviours and AUD, as well as related metabolic traits. We confirm the validity of our genetic instrument for FGF21 by demonstrating its associations with liver fat and liver cirrhosis as positive control analyses. Our findings generate important insights that support the clinical development of FGF21 analogues for the treatment of AUD.

## Methods

### Study overview

For the first step in this study, a robust genetic instrument for FGF21 agonism was identified by considering two potential candidates and investigating their Mendelian randomization estimates with the positive control outcomes of liver fat and liver cirrhosis risk. FGF21 perturbation was subsequently instrumented using *FGF21* single-nucleotide polymorphism (SNP) rs739320. This missense variant results in a leucine to proline amino acid change in position 174, and has been shown to mimic the clinical effects of FGF21 analogues, including lowering liver fat percentage and cirrhosis risk.^10–12^ This SNP is also the lead genome-wide significant signal at the *FGF21* gene in the largest genome-wide association study (GWAS) of cirrhosis.^11^

Summary data unweighted Mendelian randomization analyses were performed to investigate associations between genetically instrumented FGF21 agonism and outcome traits. Primary outcomes of interest were alcohol consumption, problematic alcohol use (PAU) and AUD. Related behavioural, cardiometabolic and renal biomarkers and outcomes are investigated as secondary outcomes. As a sensitivity analysis, statistical colocalization analyses were performed to evaluate whether Mendelian randomization evidence may be biased by confounding variants in linkage disequilibrium to rs739320. An overview of the study design is provided in **Figure 1**.

**Figure 1.**
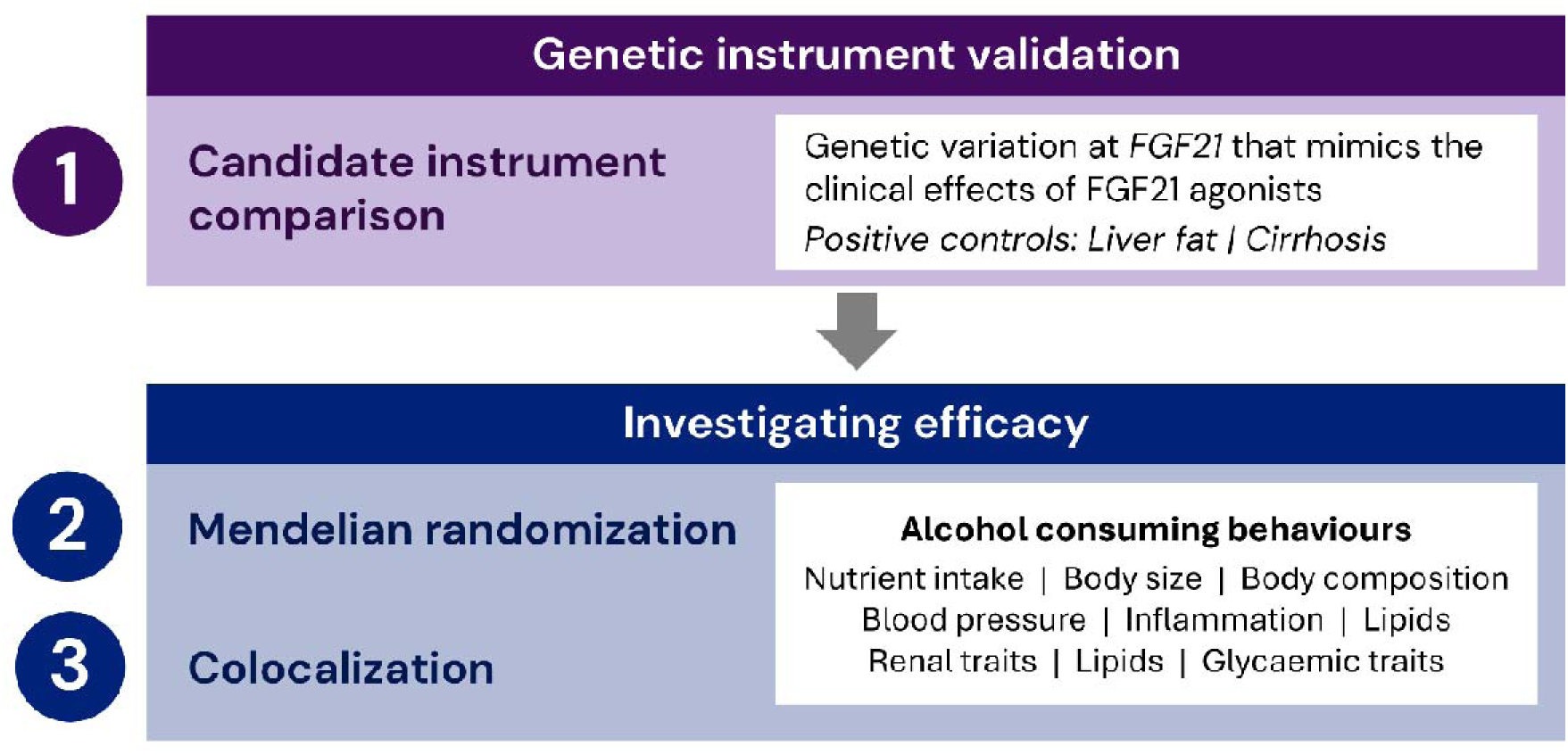
Study design.

### Data sources

Genetic association estimates for circulating FGF21 levels,^13^ magnetic resonance imaging (MRI)-derived liver fat percentage,^10^ cirrhosis liability,^12^ proportional macronutrient intake,^14^ alcohol consuming behaviour^15,16^ and substance use, ^17^ body size^18^, composition^19,20^ and bone mineral density,^21^ blood pressure,^22^ biomarkers of inflammation,^23,24^ renal function, ^25–28^ chronic kidney disease risk, ^17^ blood lipids^29^ and glycaemic traits^30^ were obtained from published studies.

All included studies had obtained the requisite ethical approval and participant consent for summary data distribution. For traits studied in our previous work,^31^ we leveraged updated (i.e., recent GWASs using data overlapping with those used before) or independent data where possible. **Table 1** provides an overview of all data sources, including a citation to their original publications wherein additional information can be accessed.

**Table 1.**
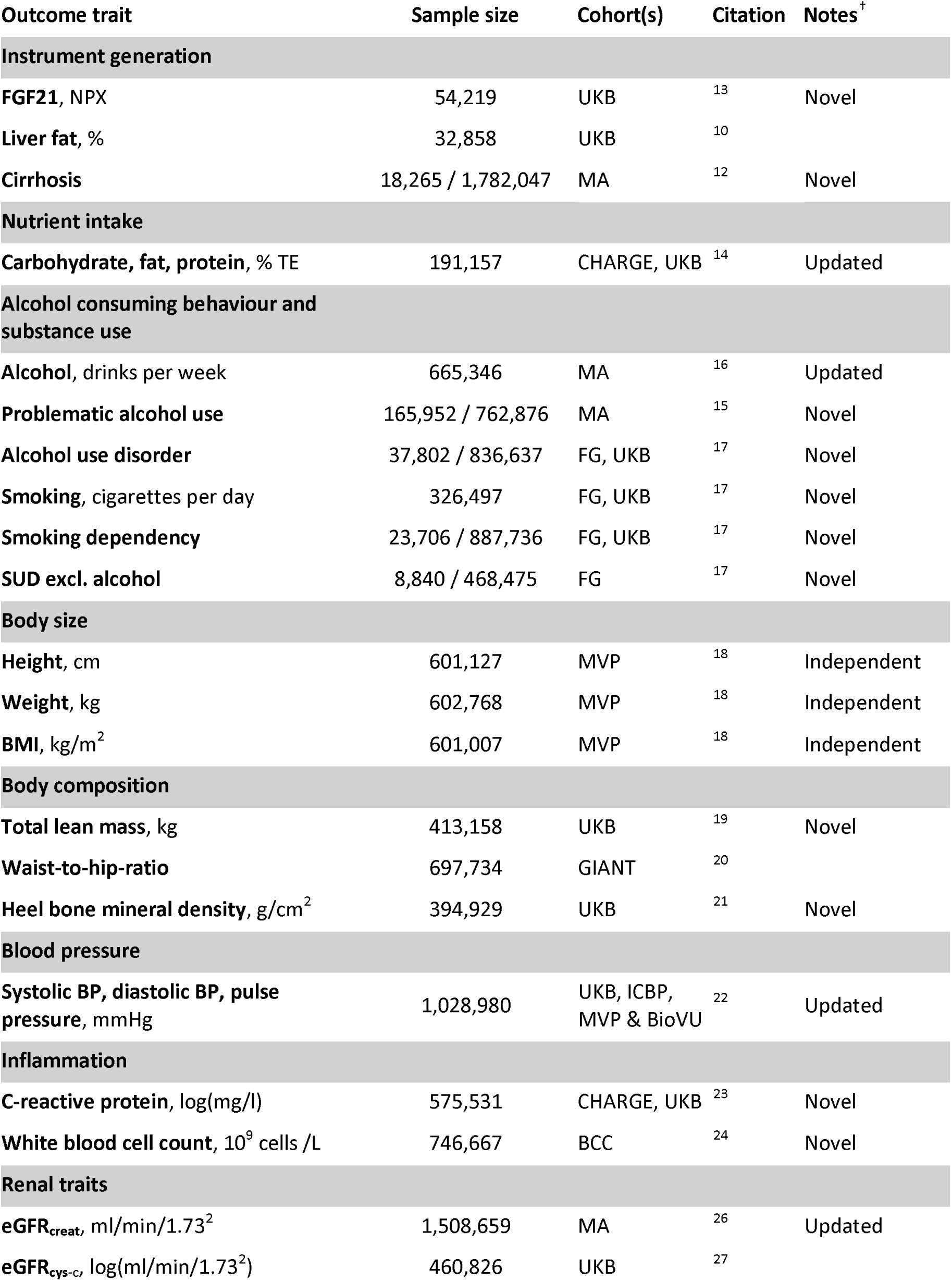
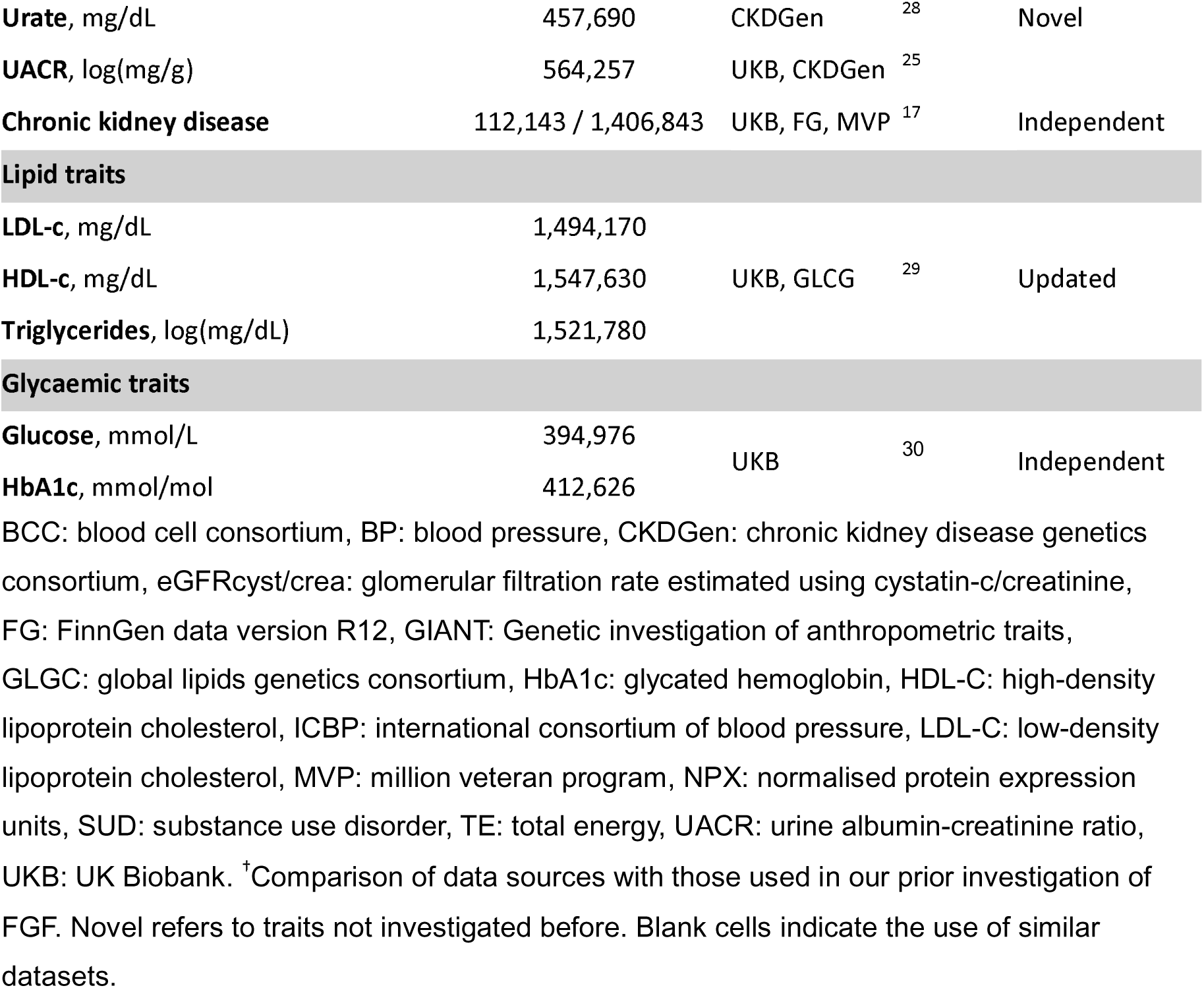
Data sources.

### Selection of the genetic instrument

We considered two candidate genetic instruments for FGF21 agonism. The first is an intronic-cis protein quantitative trait locus (cis-pQTL) variant that represents the strongest genetic association signal at the *FGF21* locus for circulating FGF21 levels (rs838131; 0.12±0.006 higher normalized protein expression (NPX) units of FGF21 per A allele; p=1x10^-88^).^13^ The other is the common allele *FGF21* L174P missense variant (rs739320) that represents the strongest genetic association signal at the *FGF21* locus for liver fat content (**Figure 2**) and cirrhosis risk.^11^ These variants are in weak linkage disequilibrium, with an R^2^ of approximately 0.5 in European cohort reference data.^32^

**Figure 2.**
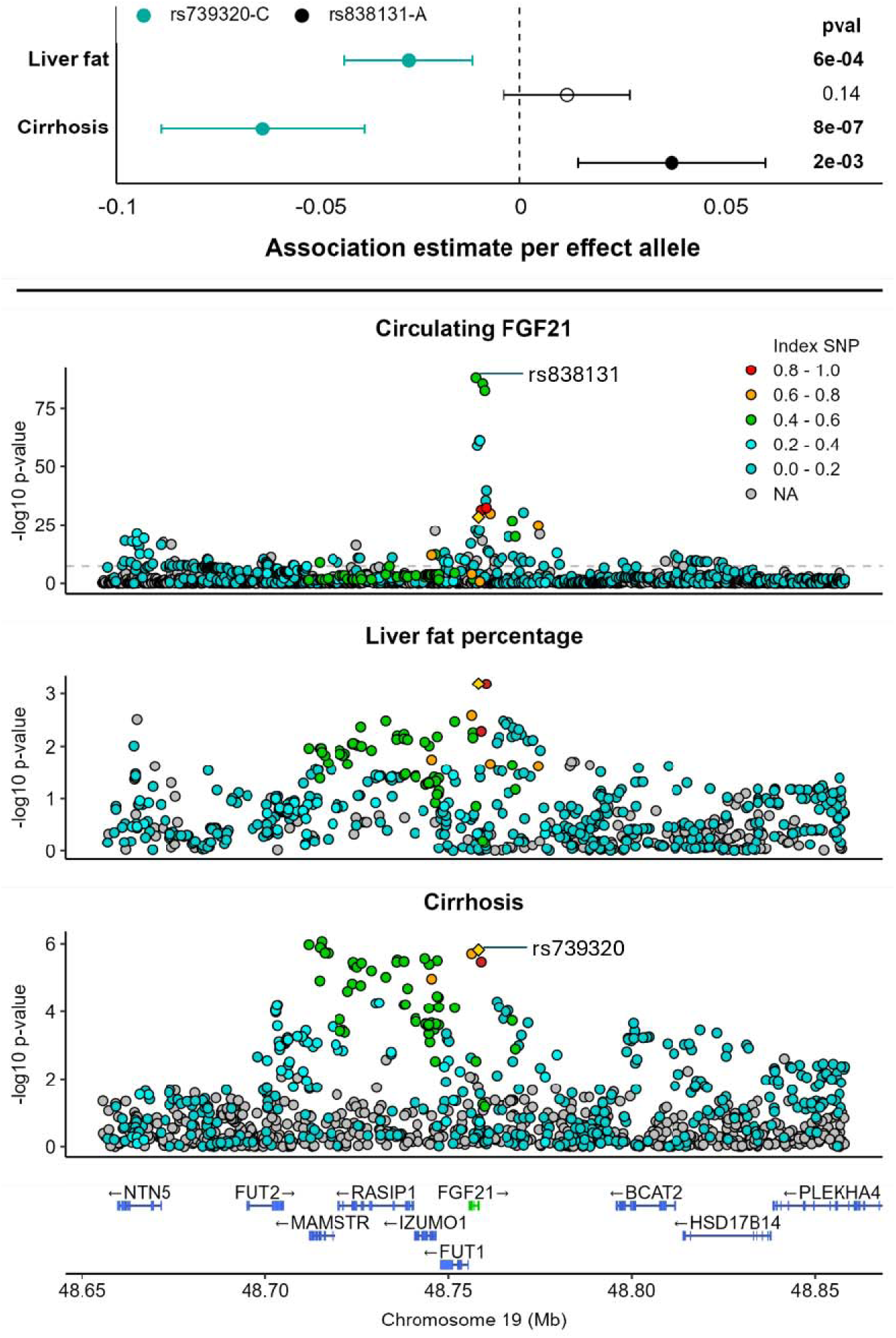
Genetic association evidence used to inform instrument selection. Top panel: Comparative genetic association estimates for rs838131-C and rs739320-C with liver fat percentage and cirrhosis risk. Bottom panel: Locus plot depicting associations between genetic variants in and around *FGF21* for circulating FGF21 levels, liver fat percentage and cirrhosis risk. The yellow triangle represents rs739320. Pairwise correlation coefficients (r^2^) with rs739320 is indicated by the colour of each data point.

Instrument validity was investigated by comparing associations of these two candidates across outcome traits known to be affected by treatment with FGF21 analogues in clinical trial settings, namely liver fat and liver cirrhosis.^4^

### Unweighted Mendelian randomization

Unweighted Mendelian randomization was performed, considering association estimates per effect allele of the variant selected as the instrument. Mendelian randomization estimates are provided as betas and 95 % confidence intervals (95% CI) per additional copy of the rs739320 C allele. The C allele is the major allele of rs739320 that is associated with lower liver fat percentage and cirrhosis risk.

Betas represent log-odds for binary outcomes, and standard deviation (SD) unit difference (95 % CI) for continuous outcomes. Where original data sources reported unstandardised association estimates, adjustments were applied based on reported trait SDs (systolic blood pressure: 19mmHg, diastolic blood pressure: 11mmHg, pulse pressure: 13.7mmHg, carbohydrate intake: 8.6%, fat intake: 6.9, protein intake: 3.5%, log-transformed cystatin C-based estimated glomerular filtration rate [eGFR]: 0.2ml/min/1.73^2^). Unadjusted p-values are presented throughout. For primary outcome traits, associations with p-values lower 0.017 are considered statistically significant, after applying a Bonferroni correction to account for multiple testing of three outcomes.

### Colocalization

We conducted colocalization analyses to investigate whether the lead genetic association signal at the *FGF21* region (*FGF21* ±100kb) for liver cirrhosis risk,^12^ is the same as that of other outcomes of interest. Aside from instrument selection, these analyses were only performed on outcomes that were statistically significantly associated with instrumented FGF21 agonism, to serve as an indicator of the probability that this evidence could be attributable to confounding through variants that are correlated to our genetic instrument due to linkage disequilibrium. Colocalization analyses were performed on 369 variants that were represented in all considered datasets using the ‘Coloc’ package (v5.2.3) set to its default priors.^33,34^ Output comprises posterior probabilities (PP) for five competing hypotheses. The first three are that causal genetic predictors exist for neither (PPH0) or one of the two traits (PPH1 for trait 1 and PPH2 for trait 2). The third and fourth hypotheses are that causal genetic predictors exist for both traits and that these are distinct (PPH3) or overlapping (PPH4).

## Results

### Instrument validation

We observed that Mendelian randomization analysis considering FGF21 agonism instrumented through the lead pQTL, rs838131, contradicted clinical trial findings, as the plasma FGF21-increasing A allele did not significantly associate with liver fat percentage, and associated with higher risk of liver cirrhosis. In contrast, the C allele of rs739320 was strongly associated with both lower liver fat and lower cirrhosis risk (odds ratio=0.94, 95% CI=0.91 to 0.96) (**Figure 2**).

Investigating the wider genomic region, *FGF21*±100kb, we confirmed that the genetic predictors of affinity-based measures of circulating FGF21 were not the same as those for liver cirrhosis liability (PPH3=0.88). It is therefore likely that FGF21 pQTLs may be confounded by the effects of neighbouring correlated variants in linkage disequilibrium, such as those affecting expression of the FUT2 gene.^35^

Overall, considering that *FGF21*-rs739320 is a functionally annotated variant that mimics the effect of FGF21 analogues observed in clinical trials, all further Mendelian randomization analyses for the effects of FGF21 were performed using this variant.

### Predicted effects of FGF21 agonism on alcohol-consuming behaviours

We observed consistent associations between instrumented FGF21 perturbation and behavioural traits related to alcohol use. The C allele of rs739320, which is associated with lower liver fat percentage and cirrhosis risk, was also associated with lower alcohol consumption and lower risk of problematic alcohol use and AUD (**Figure 3**). Colocalization analyses confirmed the robustness of these findings. The potential effect of FGF21 agonism on addictive behavioural traits appears to be specific to alcohol consumption, as associations were not observed with cigarette smoking behaviour, risk of smoking dependency or risk of substance use disorder excluding alcohol (**Supplemental Table 1**).

**Figure 3.**
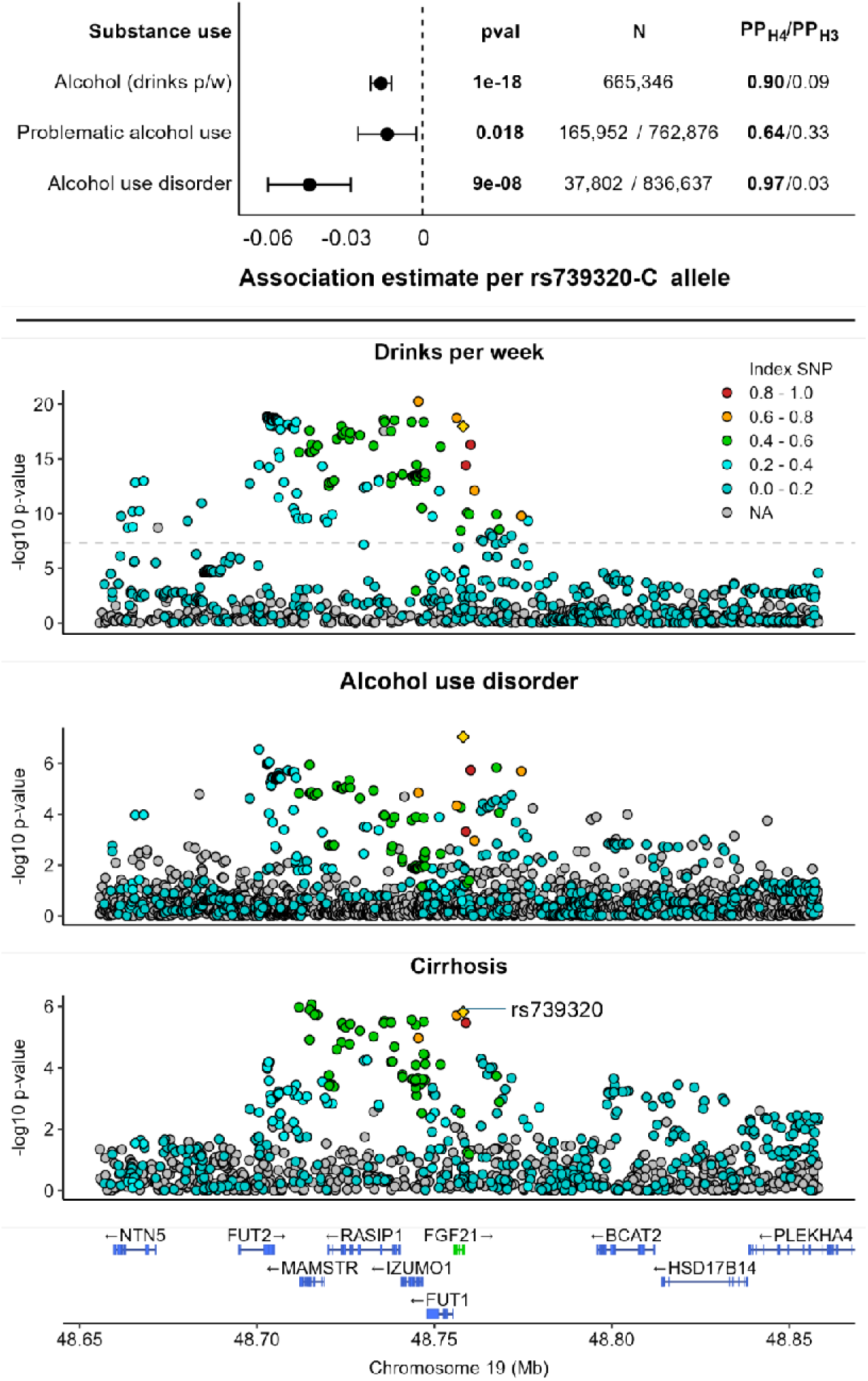
Genetic association evidence implicating FGF21 in alcohol consuming behaviours. Top panel: Unweighted Mendelian randomization estimates alongside unadjusted association p-values, sample size information and colocalization results. PPH4: Posterior probability that genetic predictors are shared with those for cirrhosis. PPH3: Posterior probability that genetic predictors are independent of those for cirrhosis. Bottom panel: Locus plot depicting associations between genetic variants in and around *FGF21* for alcohol consumption (drinks per week),risk of alcohol use disorder and cirrhosis risk. The yellow triangle represents rs739320. Pairwise correlation coefficients (r^2^) with rs739320 is indicated by the colour of each data point.

### Predicted effects of FGF21 agonism on cardiometabolic risk factors

Results for cardiometabolic and other traits are summarised in **Figure 4**. Instrumented FGF21 agonism was associated with higher body size (height, weight and body mass index), but lower central adiposity as represented by waist-to-hip ratio (WHR). No association was observed with bone mineral density. Prior reported associations of genetically predicted FGF21 agonism with lower relative intake of carbohydrates and higher relative protein and fat intake were replicated.^31^ Similarly, we replicated protective associations of instrumented FGF21 agonism with blood pressure traits, blood lipids, renal function, and chronic kidney disease risk (odds ratio: 0.98, 95% CI: 0.97 to 0.99; p= 0.004),^31^ but observed a potential adverse association with glycated haemoglobin and no association with blood glucose. Finally, we observed lower white blood cell counts and C-reactive protein levels with genetically predicted FGF21 agonism.

**Figure 4.**
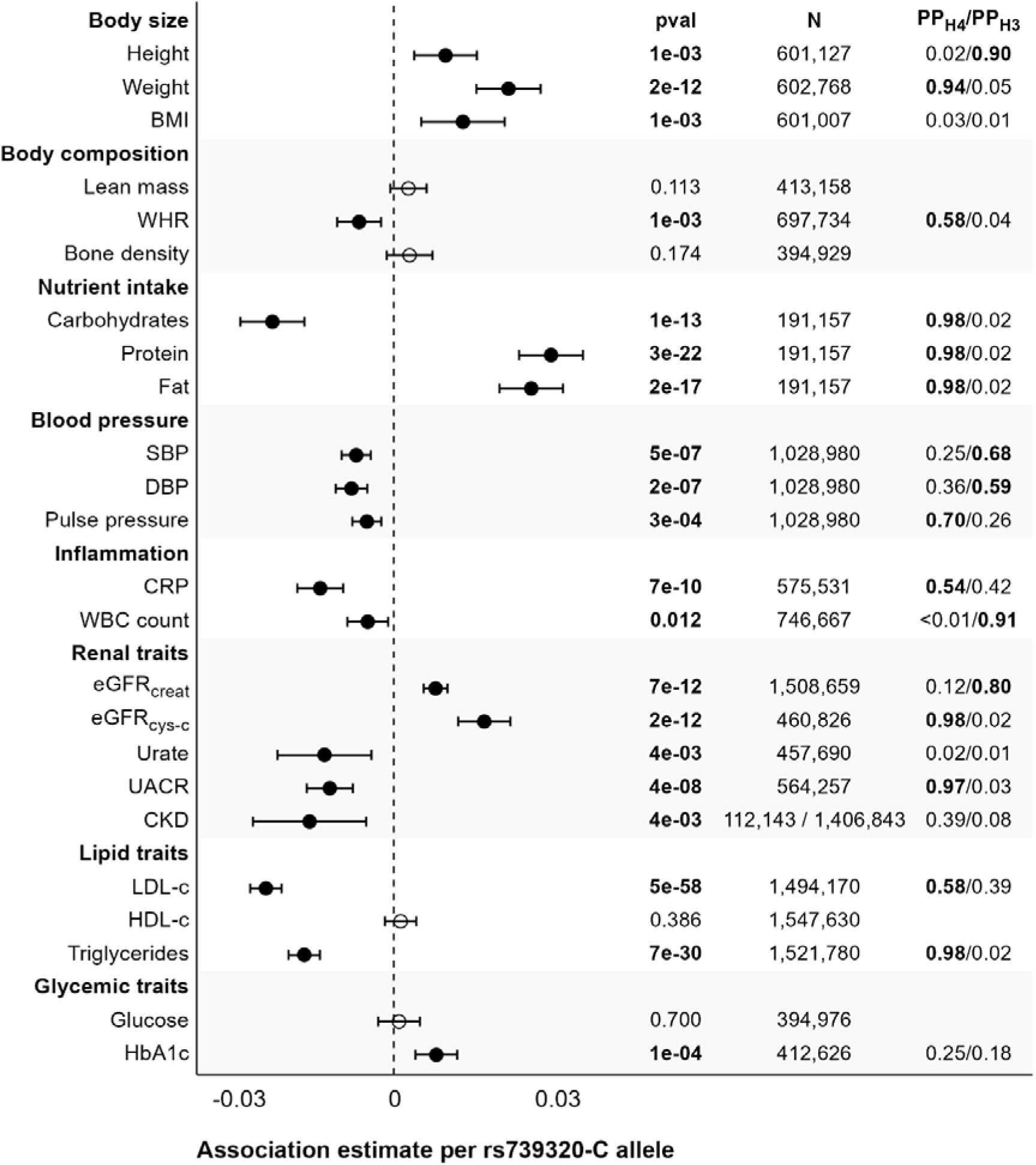
Genetic associations with biomarkers and risk factors of cardiometabolic and kidney disease risk. Association estimates represent change per additional rs739320 C allele as an instrument for FGF21 agonism. Association p-values, sample size information and colocalization results are provided as well. PPH4: Posterior probability that genetic predictors are shared with those for cirrhosis. PPH3: Posterior probability that genetic predictors are independent of those for cirrhosis.

Leveraging colocalization analyses we report strong evidence to support that the genetic predictors of cirrhosis risk at the FGF21 locus overlap with those for body weight, WHR, macronutrient distribution, pulse pressure, C-reactive protein, cystatin C-based eGFR, urinary albumin-creatinine ratio and low-density lipoprotein cholesterol, and triglycerides. Data for CKD and serum urate and glycated haemoglobin did not have sufficient statistical power for colocalization analyses.

## Discussion

### Main findings

In this study, we performed Mendelian randomization analyses leveraging a missense variant in the *FGF21* gene that strongly associates with the known clinical effects of FGF21 analogues, namely lowering of circulating triglycerides, liver fat and risk of cirrhosis. We further show that while our chosen instrument does recapitulate the known clinical effects of FGF21 agonism, this is not observed with the lead cis-pQTL for FGF21. The discrepancy may be attributable to genetic confounding through effects of variants in linkage disequilibrium, such as those affecting expression of the neighbouring FUT2 gene.^35^

Performing Mendelian randomization analyses, we provide consistent evidence to support favourable effects of FGF21 agonism on alcohol consumption behaviours, including alcoholic drinks per week, problematic alcohol use and AUD. Colocalization analysis further supported that the findings for alcoholic drinks per week and AUD are unlikely to be due to confounding from variants in linkage disequilibrium.

Considering metabolic and dietary behaviours more generally, we provide Mendelian randomization and colocalization evidence supporting effects of FGF21 on increasing growth (including both height and weight), improving body weight distribution (lowering WHR), lowering carbohydrate intake and increasing protein and fat intake, lowering C-reactive protein, improving kidney function (increasing eGFR and lowering urinary protein-creatinine ratio), and lowering LDL-C levels. We find no genetic evidence supporting effects of FGF21 (favourable or unfavourable) on bone mineral density.

### Findings in context

AUD carries a significant societal burden, affecting approximately 30 million individuals globally and accounting for £249 billion (USD) annually in social and healthcare costs.^36^ Few treatments have been approved for AUD,^9^ with the available medications having limited efficacy,^37^ and being underused.^38^ While emerging evidence supports effects of glucagon-like peptide 1 analogues (GLP1) for reducing alcohol consumption,^39^ this is proposed to have a complementary pharmacological mechanism to FGF21,^6,40^ The diverse mechanism of action of FGF21 may offer considerable and additive benefit to currently available and exploratory mechanisms of action in AUD including approved opioid receptor antagonists and the emerging GLP-1 class. ^41^

Previous Mendelian randomization analyses have supported favourable effects of FGF21 overlapping with those investigated in our current work.^31^ However, this current study makes several notable advances. Firstly, we explore a greater number of traits related to alcohol intake behaviours, to increase conviction on the potential efficacy of FGF21 analogues for AUD. Secondly, studies with larger sample sizes are considered, to improve statistical power. Thirdly, we perform colocalization analyses to help exclude the possibility of genetic confounding explaining our observed Mendelian randomization findings.

### Implications for further research

Clinical drug development has a failure rate of approximately 90%, largely due to the pre-clinical evidence typically being drawn from animal experiments and traditional epidemiological studies, which are limited their human relevance and ability to draw causal inferences.^42,43^ Drug targets supported by human genetic data are more than twice as likely to advance to patient care,^44^ and such genetic insights are becoming increasingly accessible as the availability of large-scale genetic association datasets and genotyped biobanks grows. The findings of this study represent causal human evidence to support effects of FGF21 agonism on reducing alcohol consumption and risk of AUD, alongside a plethora of favourable metabolic effects, which complement the beneficial effects of FGF21 analogues on lowering triglycerides, liver fat and liver cirrhosis risk that have already been observed clinically.^2,4^

Our current findings also triangulate with the existing animal and observational data implicating FGF21 in regulating alcohol consumption as well as the effects of alcohol. Overexpression of FGF21 in transgenic mice results in lower alcohol preference,^45^ and pharmacological administration of FGF21 analogues reduces alcohol consumption in mice and non-human primate models.^46^ There is also human epidemiological data linking FGF21 to AUD,^47^ although directionality of effect is difficult to infer without randomization of an FGF21 intervention. Our current Mendelian randomization findings strengthen the existing data by adding a causal framework in humans. Taken together, the totality of evidence strongly implicates FGF21 agonism as an efficacious therapeutic strategy for treating AUD.

### Strengths and limitations

Our work has several strengths. Firstly, the Mendelian randomization paradigm infers causality in humans, thus supporting efficacy of FGF21 analogues for treating AUD in the clinic. Secondly, we complement the Mendelian randomization evidence with statistical colocalization, to support that the findings are unlikely to be attributable to genetic confounding through neighbouring variants in linkage disequilibrium, Third, we consider a range of complementary traits related to alcohol consumption behaviours from distinct data sources, which all support similar conclusions to strengthen the body of genetic evidence. Finally, we leverage a clinically validated instrument, in contrast to the cis-pQTL for FGF21, which does not recapitulate known effects of FGF21 analogues when applied in the Mendelian randomization framework.

Our work also has limitations. Firstly, the analyses were performed using data from largely European populations and so may not necessarily be extrapolated to other population groups. Secondly, the insights are limited by the available genetic association data. As such there may be the possibility of false negative findings for underpowered outcomes. Thirdly, absence of colocalization evidence may be attributable to low statistical power or confounding signals which may mask true shared causality, thus limiting interpretability. Finally, these genetic analyses consider the effects of small lifelong variation in FGF21 agonism and may not reliably mimic the effect of a clinical intervention at a discrete timepoint in adult life. Caution should therefore be taken in extrapolating Mendelian randomization findings to the clinic.

## Conclusion

The study provides robust causal human evidence to support favourable effects of FGF21 agonism in the treatment of AUD and related outcomes. Further clinical study investigating FGF21 as a therapeutic target for this unmet clinical need and massive societal burden is duly warranted.

## Data Availability

All data produced in the present work are contained in the manuscript.

## Acknowledgements

The authors thank the participants and investigators of the cohort studies incorporated in our work. Publications from which summary-level data were acquired are cited throughout this manuscript.

## Sources of Funding

This work was supported by Apollo Therapeutics Limited. M.V. is supported by National Institutes of Health and National Institute of Diabetes and Digestive and Kidney Diseases grant R01-DK134575. S.C.L. is supported by the Swedish Research Council (Vetenskapsrådet; grant number 2019LJ−LJ00977), the Swedish Heart-Lung Foundation (Hjärt-Lungfonden; grant number 20210351), and the Swedish Cancer Society (Cancerfonden).

## Disclosures

H.T.C., S.H. and D.G. are employees of Sequoia Genetics, a private research and development consultancy that works with investors, pharma and biotech in leveraging human genetic data to inform drug discovery and development. Sequoia Genetics received funding from Apollo Therapeutics to undertake this work. R.B., R.G. and R.M. are employees and shareholders of Apollo Therapeutics. D.G. is a Special Advisor to Apollo Therapeutics and has financial interests in several private biotechnology companies. M.V. and S.C.L. have no relevant disclosures.

## Supplemental Tables

**Supplemental Table 1.** Genetic associations with smoking behaviours and substance use risk not including alcohol.

| Outcome | Beta | SE | p-value |
| --- | --- | --- | --- |
| **Smoking**, cigarettes per day | 0.004 | 0.003 | 0.152 |
| **Smoking dependency**, log-odds | -0.005 | 0.010 | 0.584 |
| **SUD excl. alcohol**, log-odds | -0.009 | 0.017 | 0.609 |

SUD: substance use disorder. Association estimates represent change per additional rs739320 C allele as an instrument for FGF21 agonism

## Notes

### Author Declarations

All included studies had obtained the requisite ethical approval and participant consent for summary data distribution.

